# Use of serology to assess the probability of public health action needed for trachoma in coastal Ecuador

**DOI:** 10.64898/2026.02.18.26346552

**Authors:** Everlyn Kamau, Lesly Simbaña Vivanco, Stuart Torres Ayala, Nikolina Walas, Gretchen Cooley, Chabier Coleman, E. Brook Goodhew, Diana L. Martin, Hadley Burroughs, Manuel Calvopiña, William Cevallos, Sandra Vivero, Victoria Nipaz, Josefina Coloma, Gwenyth O. Lee, Gabriel Trueba, Joseph N.S. Eisenberg, Karen Levy, Benjamin F. Arnold

## Abstract

We evaluated the probability of need for public health action against trachoma in Esmeraldas province, Ecuador. Compared to global references, seroconversion rates to *Chlamydia trachomatis* Pgp3 in children suggest high probability of action needed in rural villages (91%) and lower in more urban areas (32%), motivating further trachoma assessment.

## Background

Trachoma, a blinding disease caused by infection with ocular strains of *Chlamydia trachomatis (C. trachomatis)*, is targeted for elimination as a public health problem by the World Health Organization (WHO) defined as trachomatous inflammation–follicular (TF) in ages 1–9 years being <5% in each formerly endemic district, trachomatous trichiasis (TT) unknown to the health system among ages ≥15 years being <0.2% in each formerly endemic district and there being a system in place to manage TT [1]. Public-health interventions for trachoma include <u>s</u>urgery for trichiasis; <u>a</u>ntibiotic mass drug administration (MDA); and programs to support facial cleanliness and <u>e</u>nvironmental improvement (SAFE) [2].

*C. trachomatis* anti-Pgp3 IgG antibodies are a sensitive marker of past infection, making Pgp3 suitable for monitoring populations post-antibiotic MDA [3]. When measured in population-based surveys, Pgp3 can be translated into measures of transmission intensity [4], and a global analysis developed methods to estimate the probability of the need for public health action based on the Pgp3 seroconversion rate (SCR) [5].

The highest prevalence of trachoma is found in sub-Saharan Africa [2]. Trachoma remains a public health concern in vulnerable, remote areas in South America, notably Brazil, Colombia, Peru, and potentially Bolivia, Ecuador, and Venezuela, with ongoing Pan American Health Organization (PAHO)-supported efforts using the SAFE strategy [6]. Little is known about trachoma in Ecuador. We recently identified high levels of incident seroconversion to Pgp3 in a birth cohort in Esmeraldas Province in coastal Ecuador [7]. Here, we augment birth cohort samples to include older children from villages in the same region to enable estimates of the Pgp3 SCR and compare them with global distributions to estimate the probability of need for public health action against trachoma.

## Methods

### Study populations

We included samples from two studies. The ECoMiD longitudinal birth cohort in Esmeraldas Province, which enrolled pregnant mothers from 2019 through 2022 along an urban-rural gradient that included Esmeraldas city (urban), Borbón (commercial center), four communities accessible by road, and four communities accessible via the Santiago and Onzole rivers (**Supplementary Figure 1**) [8]. Dried blood spots were collected between 2021 and 2024 when children were aged 6, 9, 12, 18, and 24 months [7]. We also included samples from community-based, annual serological surveillance for arbovirus transmission in Borbón plus five communities, four of which overlapped ECoMiD (Maldonado, Colon Eloy, Timbiré, Santo Domingo) plus an additional community, Santa Maria, on the Cayapas river [9]. Annual surveys were conducted between August to October and used stratified sampling, oversampling 2–14-year-olds. All serum samples collected from 2–15-year-olds in the 2021 and 2022 surveys were included in this analysis. Samples were tested for IgG to Pgp3 on the Luminex MAGPIX instrument at Universidad San Francisco de Quito [7].

We combined data from both cohorts and stratified the communities into two groups: Borbón (the commercial center for the region) and rural villages. We excluded birth cohort samples from Esmeraldas city (n=355) because the city was not included in the arbovirus surveillance study. **Supplementary material** includes details on study setting, sample testing and statistical analysis.

### Estimation of SCR and probability of public health action

SCR is currently preferred to seroprevalence to guide decision-making [5]. We used SCR to assess probability of whether *C. trachomatis* transmission is present at a level that might warrant public health action. For each group, Borbon and rural villages, we fitted a catalytic model to the age-seroprevalence data to estimate a single seroconversion rate per 100 person-years using a generalized linear model (**Supplementary Material**), assuming a constant force of infection. Then given the SCR estimates, we estimated the probability of each group falling into either “Action needed” or “Action not needed” categories [5]:

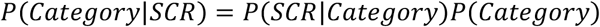

where “Action needed” are populations believed to likely experience development of disease sequelae and blindness from trachoma in the absence of effective interventions and “Action not needed” are populations believed unlikely to have sufficiently intense and sustained ocular *C. trachomatis* infections to lead to blindness, and thus no contemporary justification for population-level interventions.

The likelihood *P*(*SCR*|*Category*) was calculated as the empirical probability of the SCR estimates from Ecuador relative to SCRs in a global trachoma serology dataset derived from well characterized populations [10] (**Supplementary Material**). We considered the serological survey in coastal Ecuador a baseline survey (not necessarily post-elimination) and used an uninformative prior of category, i.e., *P*(*Category*) = 0.5, following previous analyses [5]. That is, since we identified no prior published information on trachoma epidemiology in coastal Ecuador, the communities were assumed equally likely to need trachoma-related public health action or intervention. For each group, we also calculated the probability that the SCR is above (or below) specific thresholds for elimination as a measure of confidence of the need for public health action. As no WHO guidelines on using SCR thresholds have yet been published, we considered SCR of 2 and 4 per 100 person-years as illustrative boundaries for evaluating trachoma elimination following previous analysis[5]. These values align with regions of high confidence (>90% probability) of “Action not needed” (SCR < 2) versus “Action needed” (SCR > 4) [5].

## Results

### Seroprevalence and seroconversion rates

A total of 2,806 dried blood spots from 1,243 children aged 0-15 years were analyzed for anti-Pgp3 IgG. Overall, seroprevalence was 7.0% (56/800, 95% CI 5.3% to 9.0%) in Borbon and ranged from 0% (0/38, 95% CI, 0% to 9.3%) in Colon de Onzole to 31% (17/55, 95% CI,19.1% to 44.8%) in Zancudo (**Figure 1A**). Age-specific seroprevalence rose earlier and to a higher level in rural villages compared with Borbón and groups converged by age 13 years (**Figure 1B**). Sample sizes were larger at the youngest ages through contributions from the birth cohort, with far fewer 3–5-year-olds contributing samples (**Figure 1C)**.

**Figure 1.**
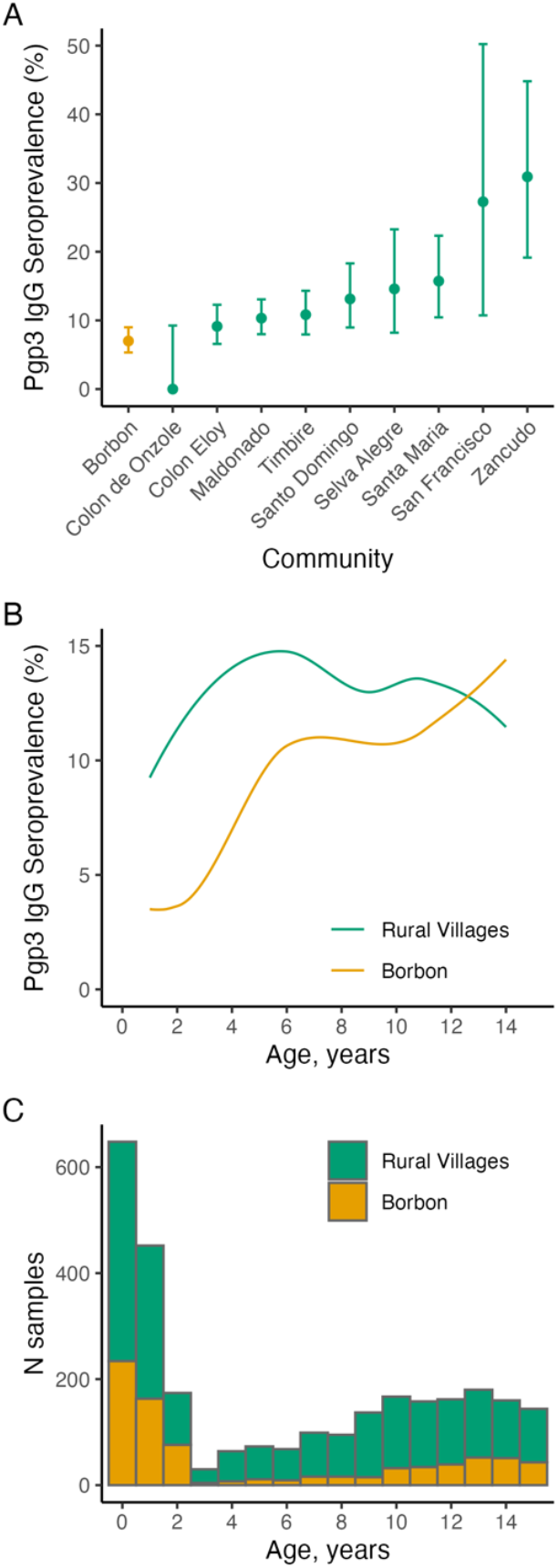
Seroprevalence to *Chlamydia trachomatis* Pgp3 in Esmeraldas province, Ecuador, 2021-2024. **A.** Seroprevalence among all children ages 0-15 years old in Borbon (commercial center) and in nine rural villages. **B**. Seroprevalence by age and community type estimated using locally weighted regression, trimmed to ages 1-14 years to reduce edge effects. **C**. Number of samples by age in years completed and community type. A total of 2,806 samples were collected from 1,243 children ages 0-15 years old. There were 790 samples from 401 children ages 1-5 years old.

Among 1–5-year-olds (401 children and 790 samples), the SCR was 4.6 (95% CI, 3.5 to 6.2) per 100 person-years. The SCR in 1-5-year-olds was 1.9 (95% CI, 0.7 to 5.5) per 100 person-years in Borbón and 5.7 (95% CI, 4.2 to 7.7) per 100 person-years in rural communities.

### Probability of need for public health action for trachoma

When compared to distributions from 34 global trachoma surveys with expert determination of the need for public health action, the probability that public health action *is* needed based on the estimated SCR was 99.3% for the rural villages and 32.0% for Borbón (**Figure 2A**). Conversely, the probability that public health action *is not* needed (1 − *P*(*action is needed*)) was 0.7% in rural and 68% in Borbón. The probability that the SCR fell below 2 per 100 person-years (i.e., *P*(*SCR* ≤ 2), a region of high confidence for no public health action needed) was 0% for the rural villages and 52.4% for Borbón (**Figure 2B**). Conversely, the probability that SCR was above 4 (i.e., *P*(*SCR* ≥ 4)) was 91.3% for the rural villages and 8.8% for Borbón (**Figure 2B**).

**Figure 2.**
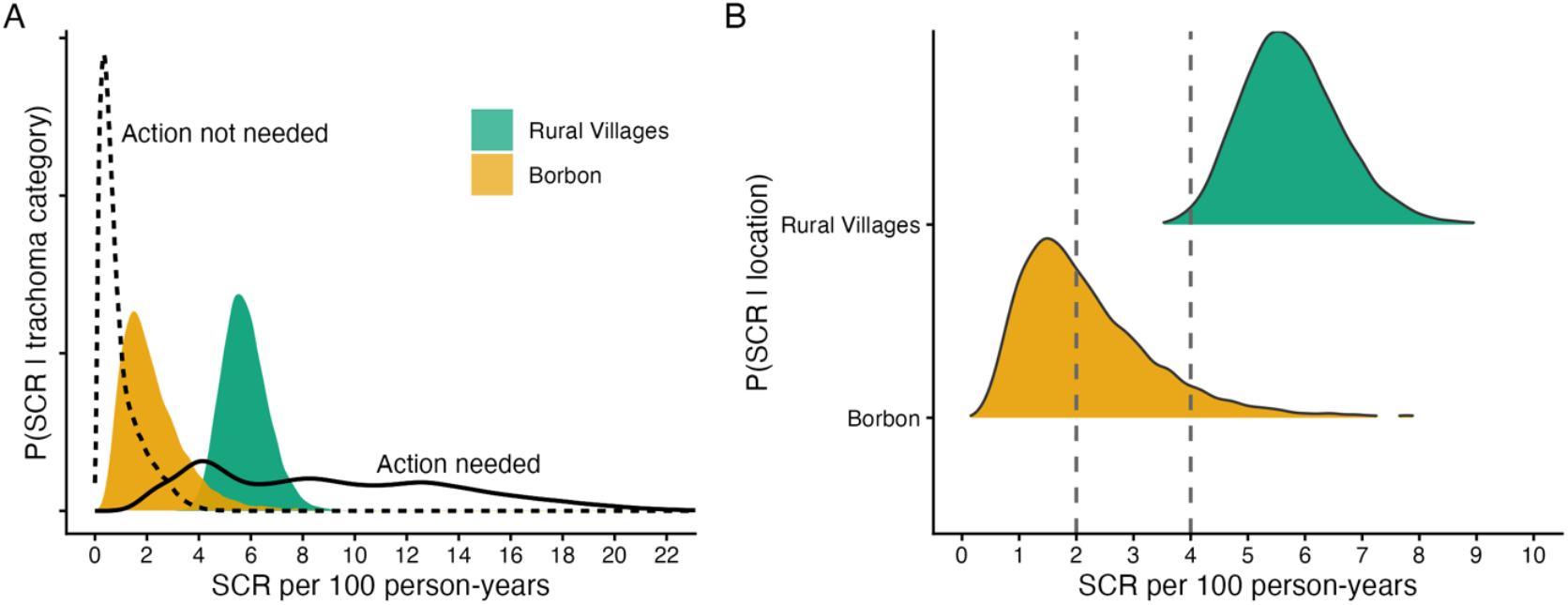
Model-based estimates of the *Chlamydia trachomatis* Pgp3 seroconversion rate (SCR) in Esmeraldas province, Ecuador compared to global trachoma serology distributions. **A.** Probability distributions of the SCR for Borbón (commercial center), and nine rural villages estimated among children 1-5 years old, super-imposed with pooled distributions of the SCR from populations classified as “Action not needed” (dashed line) and “Action needed” (solid line) regarding public health responses for trachoma control in a global analysis across a range of endemicity [5]. **B**. Probability distributions of the SCR for Borbón and rural villages from panel A compared with illustrative thresholds of 2 and 4 per 100 person-years. Below 2 has been identified as a region of >90% probability of public health Action not needed, whereas >4 is a region of ≥90% probability of public health Action needed.

## Discussion

Antibody responses from children in rural villages within the Santiago river basin suggest a high probability of public health action being needed for trachoma when compared to global reference distributions of Pgp3 SCR [10]. The SCR and seroprevalence estimates in rural villages fall within the range observed in districts with high levels of ongoing transmission requiring SAFE interventions [5], and are consistent with estimates in endemic districts considered to have persistent active trachoma in Ethiopia [11].

Efforts for trachoma elimination in the Americas, led by PAHO, have planned trachoma surveys in Ecuador, with priority surveillance in the Amazon basin [12]. Our data suggests that Esmeraldas Province should be considered as well. Although there are no formal WHO Pgp3 SCR thresholds for stopping or starting interventions, we used SCR of <2 and >4 as regions of high confidence for “Action not needed” versus “Action needed”. The results motivate additional trachoma surveillance (such as standardized baseline surveys, ophthalmologic assessments) and strengthened water, sanitation, and hygiene (WASH) interventions before initiating MDA in rural communities in the study communities. The results may warrant additional monitoring in Borbón, as the 95% CI of the SCR spans the intermediate range of 2–4 per 100 person-years.

These findings are relevant in the context of the Americas for identifying settings at risk of persistent or re-emerging transmission, particularly in rural areas characterized by limited access to safe WASH. Limitations of this analysis are that the sampling strategy did not use a population-based probability sample with many sampling clusters, as is commonly recommended for trachoma surveillance [13–15],. One implication is that children aged 3–5 years were under-represented, particularly in Borbón. Additionally, we did not characterize blood samples with respect to clinical signs of trachoma or ocular *C. trachomatis* infection. A natural next step would be to conduct appropriate baseline surveys using standardized methodologies [13–15], to ascertain presence of active trachoma and provide reference points against which subsequent progress can be measured.

WHO aims to eliminate trachoma as a public health problem by 2030. This analysis provides a generalizable example of using existing serosurveys to inform public health decision-making as we approach elimination.

## Acknowledgements

The authors are grateful to Dr. Anthony Solomon for his comments on a draft of this paper.

## Author Contributions

Following CRediT taxonomy: conceptualization (EK,BFA), data curation (LSV,STA,NW,BFA), formal analysis (EK,BFA), funding acquisition (BFA), methodology (EK,BFA), project administration (BFA), software (EK,BFA), supervision (BFA), validation (EK,BFA), visualization (EK,BFA), Writing – Original Draft Preparation (EK,BFA), Writing – Review & Editing (all authors)

## Data availability

Replication files are available through the Open Science Framework (https://osf.io/najfz/). Due to the small population of the study region, UCSF IRB determined that individual-level data that included child age (essential for the analysis), even if de-identified, would be considered potentially identifiable. Please contact the corresponding author regarding possible access to data underlying this analysis, pending appropriate IRB approval.

## Ethics

Institutional review boards at University of Washington (UW; STUDY00014270), Emory University (IRB00101202), University of California, San Francisco (21−33932), University of Michigan (HUM00140967), Universidad San Francisco de Quito (USFQ; 2017-159M, 2018−022M), and the Ecuadorian Ministry of Health (MSPCURI000253−4) reviewed and approved the study protocol. Caregivers provided informed consent and field staff obtained consent and assent before each sample collection.

## SUPPLEMENTARY MATERIAL

### Supplementary Methods

#### Study se)ng, sample testing and statistical analysis

ECoMiD is a longitudinal birth cohort study in Esmeraldas Province, on the northern coast of Ecuador ^1^. The study enrolled women ages ≥18 years old in their third trimester (37 weeks’ gestation) of pregnancy who were identified through prenatal care visits and outreach in local health clinics. The study excluded women with known, high-risk pregnancies, those who anticipated moving in the next 6 months, and those who planned to have a non-vaginal (caesarean) delivery. The study enrolled pregnant mothers from 2019 through 2022 along an urban-rural gradient Esmeraldas city (urban), Borbón (commercial center), four communities accessible by road, and four communities accessible via the Santiago and Onzole rivers (**Fig S1**). Children in the birth cohort were followed for their first two years of life, with biological samples collected every three months. The study region is predominantly lowland tropical rainforest and converted agricultural land that has undergone gradual development since road construction increased in the early 2000s, bringing increased connectivity between urban Esmeraldas city, the town of Borbón, and rural villages, which has led to changes in infectious disease transmission dynamics documented through the EcoDess research consortium ^2^.

Field staff collected dried blood spot samples from children up to five times at ages 6, 9, 12, 18, and 24 months. Dried blood spots were collected on filter paper, each with six 10 μL extensions. These were then dried, individually sealed with desiccant in plastic bags, and stored at –20°C in the field office before being shipped to the USFQ at ambient temperature and stored at -20°C.

Samples from children enrolled after Ecuador’s COVID-19 pandemic shutdowns were the focus of IgG testing, spanning the 2021-2024 period. A single 10 μL extension from each participant’s sample was eluted overnight at 4°C in a Buffer B(1X PBS pH 7.2-7.4, 0.5%casein, 0,5% PVA, 0.8% PVP, 0.3% Tween 20, 0.02%NaN3, 3 ug/ml of *E. coli* extract). Eluates were further diluted with Buffer B to a final concentration of 1:400 for use in the multiplex bead assay.

Fifty microliters of the eluted samples were incubated for 1.5 hours with microsphere beads with coupled antigens in a 45-plex assay with antigens to a broad set of enteric pathogens, vaccine preventable diseases, arboviruses, and neglected tropical diseases, including Pgp3 and Ct694 (*C. trachomatis*).The concentration used was 1250 beads per well, diluted in assay buffer (1X PBS, 0.5% BSA, 0.05% Tween 20, 0.02% NaN3).

Samples were incubated for 45 minutes with a mixture of biotinylated mouse anti-human IgG (50ng/well) and IgG4 (40ng/well) from Southern Biotech, Birmingham, AL, USA. This was followed by a 30-minute incubation with R-phycoerythrin conjugated to streptavidin (250μg/well) from Invitrogen, Waltham, MA, USA, serving as a fluorescent marker. All incubations were performed at room temperature with agitation, with triple washes using PBST buffer (PBS with 0.05% Tween) between each step. Finally, the beads were resuspended in 100μL of PBS, and the plates were stored at 4°C until they were read the next day.

Bound IgG to each antigen was measured using median fluorescence intensity minus background (MFI-bg) on the Luminex platform using a MAGPIX instrument. Plates included Buffer B-only wells as blanks, along with a negative control and two pooled positive controls at different concentrations for quality assurance. Final MFI-bg values were obtained by subtracting the Buffer B blank from each antigen’s MFI values. Half (11 of 22) plates were run in duplicate to quantify within- and between-plate variability in MFI-bg values. Plates were repeated if positive control monitoring targets fell outside two standard deviations of the mean in Levy-Jennings control chart.

Seropositivity cutoffs for Pgp3 (MFI=238) and Ct694 (MFI=710) were determined using external values from a receiver operator characteristic curve analysis of known positive and negative samples at the US CDC. We focused primarily on Pgp3 responses to align with elimination surveillance studies that rely primarily on Pgp3 and used Ct694 as an adjunctive measure ^3,4^.

We estimated seroprevalence as the proportion of samples positive with exact binomial (Clopper-Pearson) 95% confidence intervals. We combined data from both cohorts and stratified the communities into two groups: Borbón (the commercial center for the region) and rural villages. We excluded birth cohort samples from Esmeraldas city (n=355) because the city was not included in the arbovirus surveillance study. We estimated prevalence differences between the two groups (Borbón and rural villages) using a linear-binomial model with robust standard errors clustered at the child-level.

#### Estimation of SCR within a generalized linear model using maximum likelihood

The seroconversion rate (SCR) from a single-rate catalytic model assuming no seroreversion is closely tied to the slope of age-seroprevalence curve. The SCR is equal to the slope of the age-seroprevalence curve divided by the complement of the seroprevalence at age A = a. It can be shown that the SCR based on this model can be estimated as the exponentiated intercept from a generalized linear model with binomial error structure and a complementary log-log link:

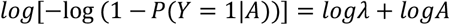

where Y represents individual-level serostatus (1: seropositive, 0: seronegative or equivocal), A is the child’s age in years, and λ is the SCR. The SCR provides an estimate of the force of infection, an epidemiological parameter that denotes the rate at which susceptible individuals in the population become infected. In this analysis, we did not allow for seroreversion in the SCR estimation.

#### Estimation of probability of public health action – ‘Action not needed’ versus ‘Action needed’

First, we downloaded posterior distributions of SCR estimates derived from a global trachoma serology dataset of well characterized populations (OSF (https://osf.io/ykjc4/) Dryad (https://datadryad.org/dataset/doi:10.5061/dryad.5qfttdzhx) ^8^. These populations had been determined by a group of experts to either require further action (Action needed) or not (Action not needed) ^3^. Therefore, the downloaded data could be labelled according to these two categories.

Then for the Ecuador data, we created a distribution of random values from the estimated log(SCR) and its standard error (SE):

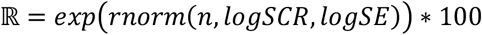

to create the likelihood that would be used in the calculation of probability as described below.

To calculate probability for each category of public health action, we used a mixture model framework applied to the distributions of SCR estimates. This approach assumes that an SCR estimate is drawn independently from a 2-component distribution of the two categories above, *k* ∈ {1, 2}. So, for each component or category (*C*_*k*_: action not needed, action needed) and SCR estimate, *x ϵ* ℝ, we computed the posterior probability, *p*(*C*_*k*_|*x*), using Bayes’ rule:

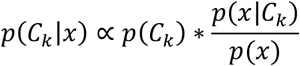

where *p*(*C*_*k*_) is the prior probability that a population is in category *C*_*k*_. In this study, *p*(*C*_*k*_) = 0.5 as we considered the Ecuador study as a baseline survey. *p*(*x*|*C*_*k*_), is the likelihood evaluated as empirical probability density function at each value *x*. In other words, the probability of *x* relative to the distribution of values comprising the category *C*_*k*_. *p*(*x*) denotes the marginal likelihood or normalizing constant for the posterior density obtained by integrating the products of *p*(*x*|*C*_*k*_) and *p*(*C*_*k*_). That is, the sum of the products of the density function and prior probability for each *k*,

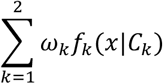

or

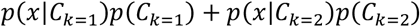

#### Estimating P(SCR ≤ c)

Using the simulated likelihood ℝ above, we calculated *P*(*SCR* ≤ *c*) as a cumulative distribution function or the empirical probability density ℝ ≤ *c*.

All analyses were done in R v4.5.0.

**Supplementary Figure 1.**
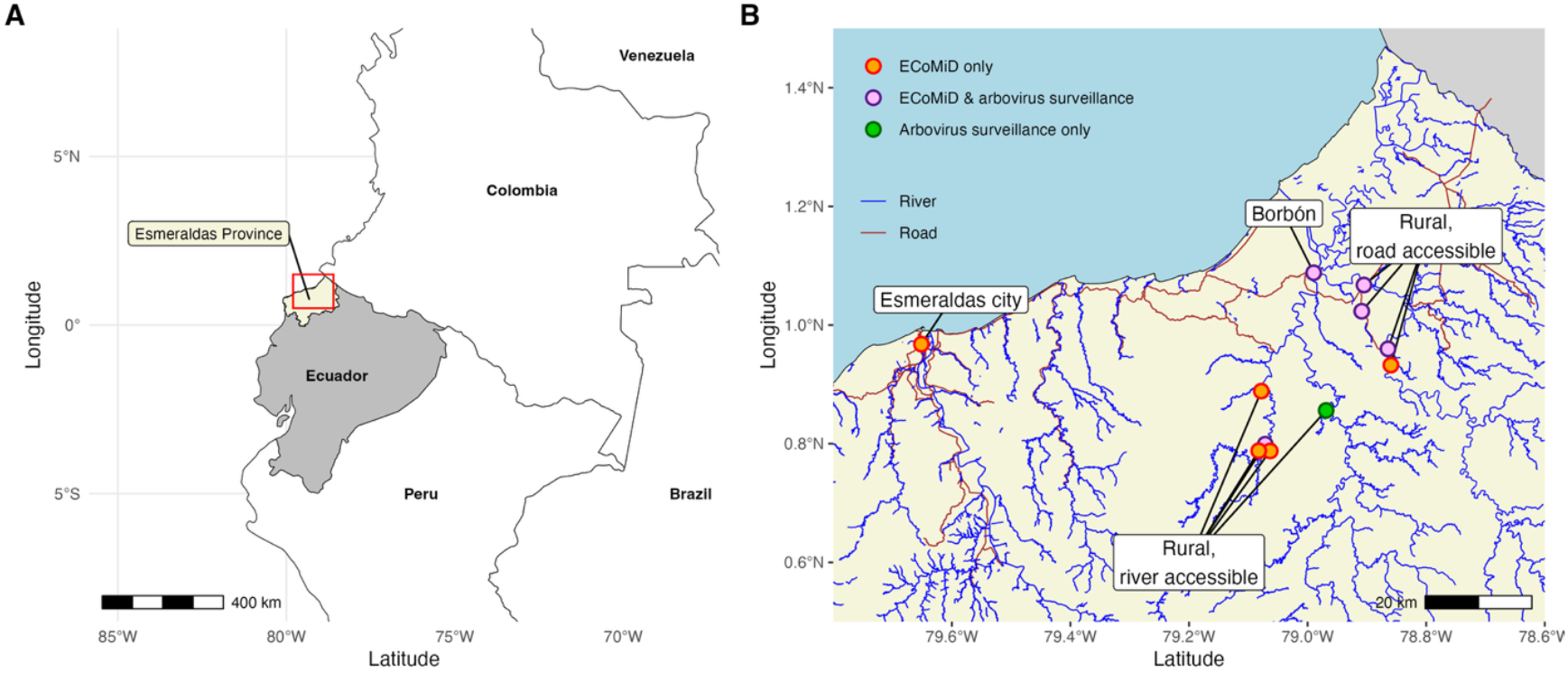
Study location in Esmeraldas province, Ecuador. **A**. Esmeraldas province is in northern coastal, Ecuador, along the border with Colombia. **B**. Children were enrolled from Esmeraldas city (population approximately 162,000), the town of Borbón (population approximately 5,000), four road accessible rural villages (populations 500 to 1000) and five river-accessible rural villages (populations 200 to 700). This study focuses on samples from Borbon and rural villages. Major rivers (blue) and major roads (brown) provided by OpenStreetMap (https://www.openstreetmap.org).

